# Cardiac Surgery during the Covid-19 Pandemic: Evidence from the first wave

**DOI:** 10.1101/2021.04.07.21254206

**Authors:** Sotiris Vandoros, Nikolaos Schizas, Alkis Apostolopoulos, Vasilios Patris, Mihalis Argiriou

## Abstract

**Background:** The Covid-19 pandemic has affected human behaviour and burdened health systems and has thus had an impact on other health outcomes.

**Objective:** This paper studies whether there was a decrease in cardiac surgery operations in Greece during the first wave of the Covid-19 pandemic.

**Data and Methods:** We used data from 7 major hospitals that geographically cover about half the country and more than half the population, including a mix of public, private, military and children’s hospitals. We used a difference-in-differences econometric approach to compare trends in cardiac surgery before and after the pandemic in 2020, to the same months in 2019, controlling for seasonality and unemployment, and using hospital fixed effects.

**Results:** We found that during the first wave of the pandemic and the associated lockdown, there were 35-56% fewer cardiac surgery operations compared to what we would have expected in the absence of the pandemic.

**Conclusions:** There was a steep decline in Cardiac surgery operations in Greece during the first wave of the Covid-19 pandemic. Possible reasons may include people not seeking medical attention to avoid the risk of catching Covid-19; fewer referrals; and working from home, thus not being exposed to a stressful work environment or commute.

## 1. Introduction

The Covid-19 pandemic has cost many lives directly, but also appears to have (positive or negative) spillover effects on other health outcomes, such as mental health (Czeisler et al 2020), cancer (Maringe et al 2020), homicides and motor vehicle collisions (Calderon-Anyosa et al 2020; Vandoros 2020a), and excess mortality in general (Vandoros 2020b). Such effects appear to extend to cardiovascular health (NYT 2020). There has been a *decrease* in the incidence of acute myocardial infarction during the early stages of the pandemic in the US (Solomon et al 2020) and a reduction in acute coronary syndrome in Italy (De Filippo at al 2020). There was a significant reduction in coronary interventions in the Veteran Affairs Healthcare System in the period from March to June 2020 compared to the previous year (Waldo et al., 2020). Similarly, recent evidence also shows that non-ST segment elevation infarctions (NSTEMI) and ST segment elevation infarctions (STEMI) decreased significantly in France during the first lockdown (Mesnier et al., 2020).

There are a number of possible reasons why the number of hospital visits relating to cardiovascular disease may be affected by the pandemic. On one hand, the worsening of economic conditions and increased unemployment may lead to more cardiovascular problems (Dupre et al 2012), and lack of physical activity or stress related to the pandemic may also play a role in that direction.

On the other hand, there might be a decrease in cardiovascular-related visits for other reasons. These may include people avoiding hospital visits to avoid catching Covid-19; health system capacity and postponed operations (in some countries, depending on the severity of the Covid-19 wave); avoiding a stressful/toxic working environment or a stressful commute to the office while working from home; and more time to cook healthier meals at home.

The objective of this study is to examine whether the number of cardiac surgery operations was affected by the first wave of the Covid-19 pandemic in Greece.

## 2. Data and Methods

We used data on monthly cardiac surgeries from major hospitals in Greece in the first six calendar months of 2019 and 2020. Data were provided by relevant departments in each hospital separately. The hospitals included in the sample (Evangelismos, Hippicratio, Attikon, Iatriko Kentro, 401 Military Hospital, Onasio, Hygeia, Aghia Sophia) are major hospitals in Athens. Evangelismos, Hippocratio, Attiko and Aghia Sophia are public; Hygeia and Iatriko Kentro are private; 401 is a Miliatry Hospital; and Onasio is a privately run public hospital. Aghia Sophia is a children’s hospital, so we analysed data with and without the inclusion of this hospital.

There were no other hospitals performing cardiac surgery operations in Athens or within a radius of 200km, thus excluding the possibility of any change in outcomes being a result of patients switching to other hospitals due to any reasons relating to the pandemic. Half the population of the country lives in Athens, but in total these hospitals cover more than half the population, as these are the closest hospitals (for cardiac surgery in particular) to people living in a number of regional units in the country.

In order to study the impact of a treatment (in this case the novel coronavirus pandemic) on an outcome, we would ideally use a control group. However, the pandemic has affected various aspects of life in every country in the world, so identifying a suitable control population or outcome seems impossible. Instead, we followed the approach of Vandoros 2020, Metcalfe et al 2011, and Powdthavwee 2019, and used the same outcome in the previous calendar year (before the Covid-19 pandemic) as a control group for the year 2020. The assumption is that in the absence of the pandemic, trends in cardiac surgery in 2020 would be the same as trends in 2019.

We used a difference-in-difference econometric approach, with the number of cardiac surgery operations as an outcome. The ‘treatment group’ is calendar year 2020; and the ‘treatment period’ includes calendar months from March to June in each calendar year (the first Covid-19 case in Greece was reported on 26 February 2021). If the Covid-19 pandemic had an effect on the number of cardiac surgeries, there would be a change in the relative trends in this outcome between 2019 and 2020 from March onwards. The empirical model is presented in equation (1):

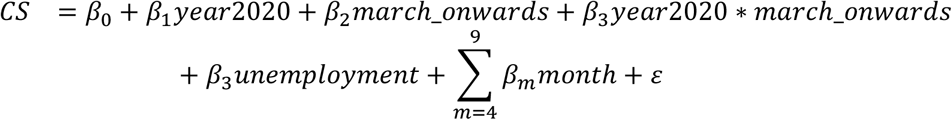

Where The dependent variable *CS* is the natural logarithm of monthly cardiac surgery operations per hospital. *year2020* is the treatment group which takes the value of 1 for calendar year 2020 and 0 therwise; *march_onwards* is the treatment period which take the value of 1 for calendar months March-June. The interaction between the two is the difference-in-differences interaction term, which is the main variable of interest. We controlled for unemployment as stress relating to the economic environment or job loss can affect cardiovascular health (Dupre et al 2012). We introduced hospital fixed effects and used month dummies to account for seasonality. Robust confidence intervals were used in all regressions.

## 3. Results

Figure 1 demonstrates trends in cardiac surgery operations in months January – June in 2019 and 2020. There is a sharp drop in cardiac surgery operations in March, April and May 2020 compared to the same months in 2019, with the number of operations returning to the expected levels in June 2020.

**Figure 1.**
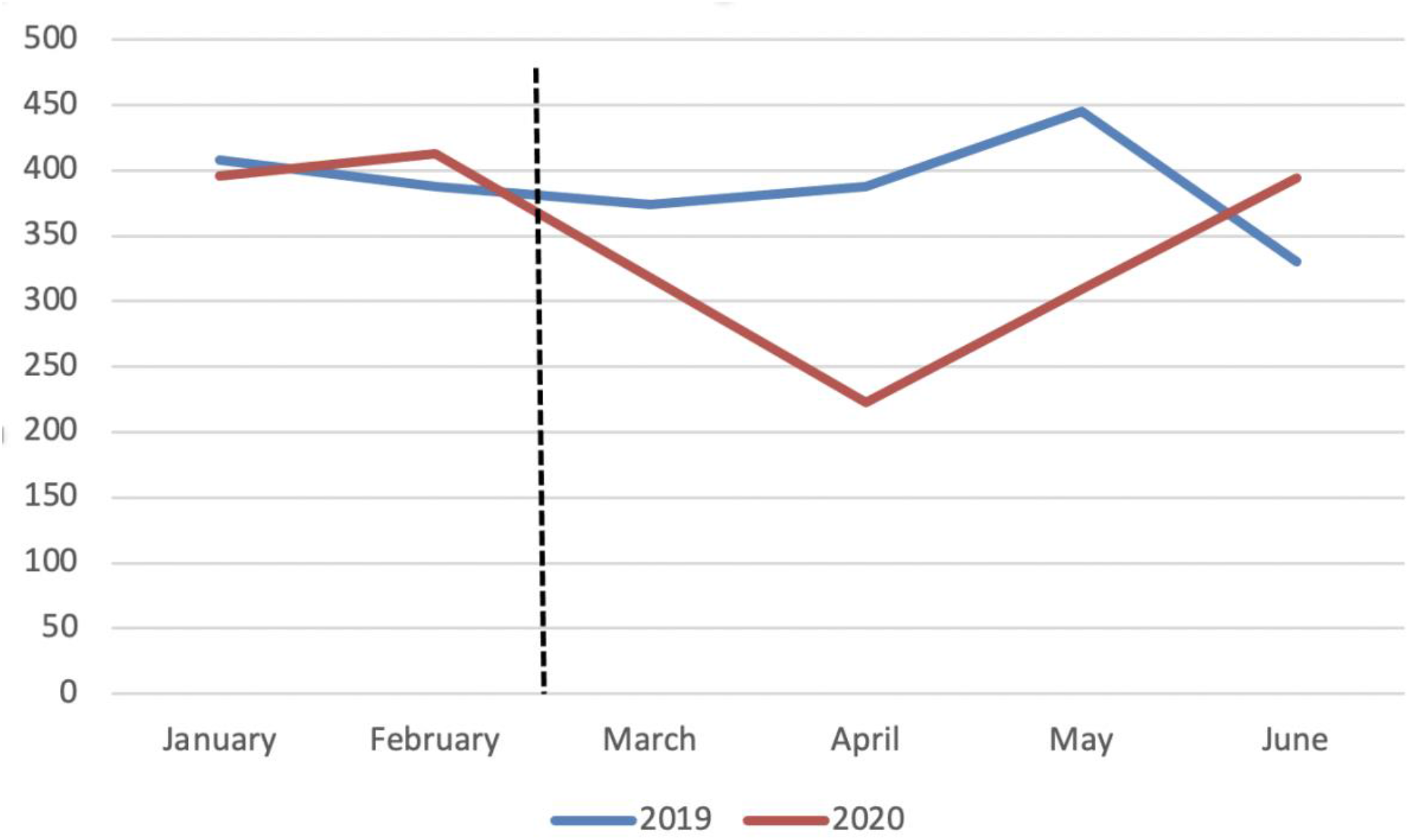
Trends in cardiac surgery operations in 7 major hospitals in Greece, 2019 and 2020. Horizontal line: First Covid-19 death and lockdown

Table 1 presents the results of the baseline econometric model. The coefficient of the DID interaction term is negative and statistically significant [Column 1 - coef:-0.35; 95% CI: −0.586 - −0.112], which suggests a 35% decrease in cardiac surgery operations compared to what would have been expected in the absence of the pandemic. Similar results hold when also controlling for seasonality [Column 2 - coef:-0.35; 95% CI: −0.576 - −0.122] as well as unemployment [Column 3 - coef:-0.53; 95% CI: −0.843 - −0.226]. Overall, our results suggest a 35 – 53% reduction in cardiac surgery-related hospitalisations.

**Table 1.**
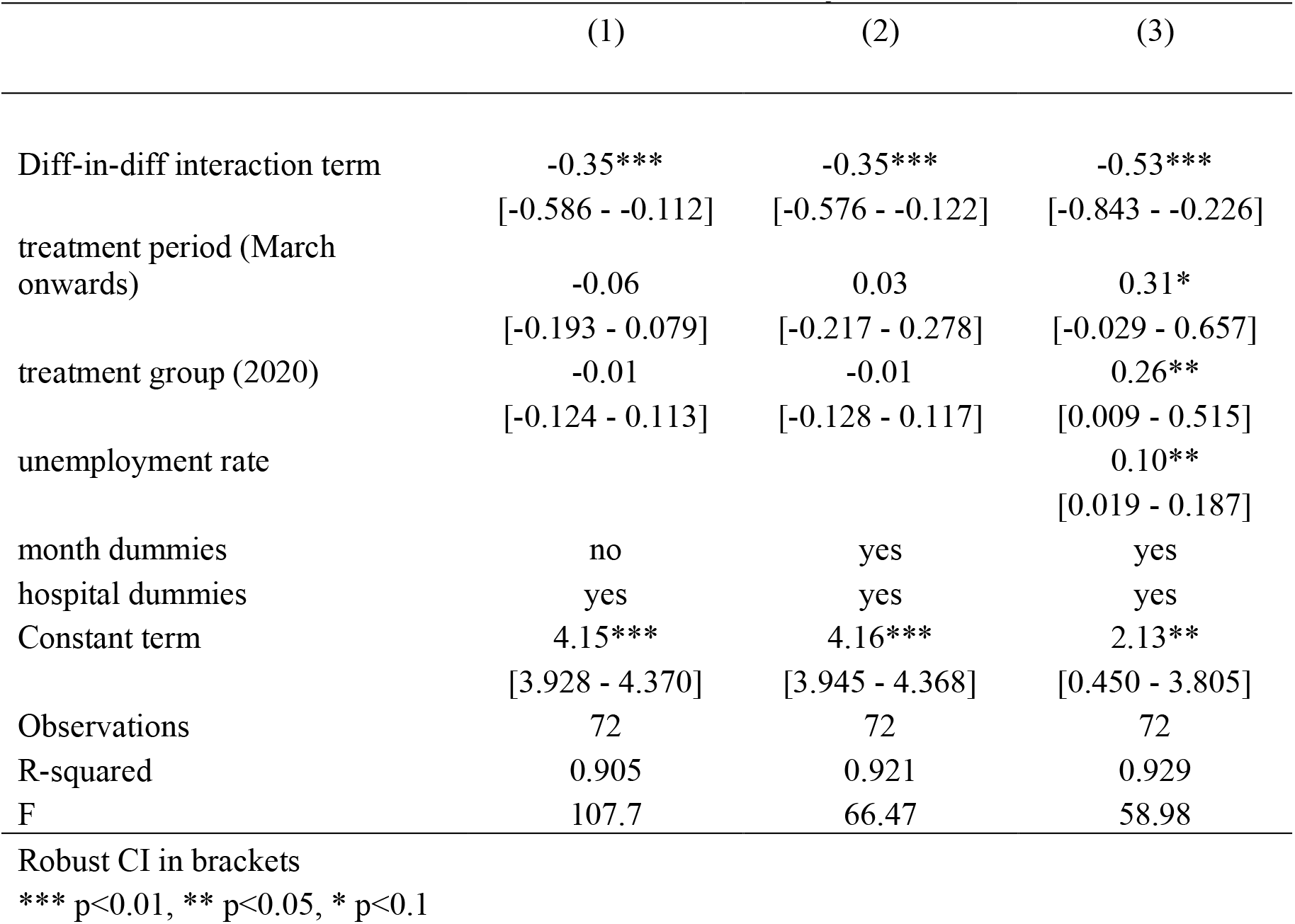
Difference-in-differences model, months January-June.

Table 2 presents the results when including the 7^th^ hospital of our sample, Aghia Sophia - a children’s hospital, in the model. Results are similar to those of the baseline model and hold the same interpretation. In the full specification (Column 3), there was a decrease in the number of cardiac surgery operations by 45% [coef:-0.45; 95% CI: −0.780 to −0.126].

**Table 2.** Difference-in-differences model, months January-June. Including Aghia Sophia hospital.

|  | (1) | (2) | (3) |
| --- | --- | --- | --- |
| Diff-in-diff interaction term | -0.30**<br>[-0.543 - -0.064] | -0.30***<br>[-0.532 - -0.075] | -0.45***<br>[-0.780 - -0.126] |
| treatment period (March onwards) | -0.10<br>[-0.243 - 0.038] | -0.02<br>[-0.260 - 0.213] | 0.21<br>[-0.155 - 0.580] |
| treatment group (2020) | -0.04<br>[-0.157 - 0.085] | -0.04<br>[-0.162 - 0.090] | 0.18<br>[-0.101 - 0.462] |
| unemployment rate |  |  | 0.08*<br>[-0.006 - 0.172] |
| month dummies | no | yes | yes |
| hospital dummies | yes | yes | yes |
| Constant | 4.18***<br>[3.958 - 4.400] | 4.20***<br>[3.986 - 4.418] | 2.56***<br>[0.770 - 4.358] |
| Observations | 78 | 78 | 78 |
| R-squared | 0.945 | 0.952 | 0.955 |
| F | 163.1 | 122.3 | 109.5 |

Robust CI in brackets. \*\*\* p<0.01, \*\* p<0.05, \* p<0.1

Finally, Table 3 presents results when restricting the sample to the first 4 calendar months, thus ending the study period at the end of the first lockdown. Once again, there are fewer cardiac surgery-related hospitalisations in March and April 2020 compared to what would have been expected in the absence of the Covid-19 pandemic. The reduction is between 53 and 56%, depending on the model specification. In the full model with all coefficients, the pandemic and lockdown were associated with a decrease in the number of cardiac surgery operations by 53% [Column 3 - coef:-0.53; 95% CI: −0.798 to −0.262].

**Table 3.** Difference-in-differences model, months January-April.

|  | (1) | (2) | (3) |
| --- | --- | --- | --- |
| Diff-in-diff interaction term | -0.56***<br>[-0.915 - -0.210] | -0.56***<br>[-0.892 - -0.233] | -0.53***<br>[-0.798 - -0.262] |
| treatment period (March onwards) | -0.04<br>[-0.173 - 0.098] | -0.21<br>[-0.461 - 0.050] | -1.13***<br>[-1.826 - -0.437] |
| treatment group (2020) | -0.01<br>[-0.139 - 0.127] | -0.01<br>[-0.142 - 0.131] | -1.69***<br>[-2.733 - -0.656] |
| unemployment rate |  |  | -0.65***<br>[-1.044 - -0.255] |
| month dummies | no | yes | yes |
| hospital dummies | yes | yes | yes |
| Constant term | 4.08***<br>[3.826 - 4.338] | 4.09***<br>[3.841 - 4.338] | 16.89***<br>[9.119 - 24.655] |
| Observations | 48 | 48 | 48 |
| R-squared | 0.911 | 0.927 | 0.948 |
| F | 49.89 | 42.23 | 62.46 |

Robust CI in brackets. \*\*\* p<0.01, \*\* p<0.05, \* p<0.1

## 4. Discussion

Using a difference-in-differences approach, we found that during the first wave of the pandemic and the associated lockdown, there were 35-56% fewer cardiac surgery operations compared to what we would have expected in the absence of the pandemic.

These findings add to existing evidence from other countries that reported a reduced number of cardiovascular-related hospital admissions during the pandemic (De Filippo et al 2020; Solomon et al 2020; NYT 2020). Our results also add to reduced hospitalisation rates for acute coronary syndromes during the first wave in Greece (Katsouras et al 2020).

There are a number of possible explanations for this reduction. First, people who were afraid of catching Covid-19 in hospital might have ignored a mild heart attack. This is an interesting case of perceived relative risk of death from Covid-19 or cardiovascular disease. Second, physician visits to GPs or cardiologists have been reduced, leading to a possible delay in diagnostic testing (stress echo, coronary angiography, scintigraphy etc). Third, there might have been a reduction in patients with STEMI (Pessoa-Amorim et al 2020). This may relate to the lockdown, which, among others, may have helped some people avoid a stressful working environment or commute and gives an opportunity to eat healthier at home.

The Ministry of Health of Greece required public hospitals to reduce the number of operations by 50% (with the exception of emergencies and oncology-related operations). However, this was not applied in practice by hospital and surgeons, as the waiting list did not increase.

In any case, we should not ignore other factors that may have put people at higher risk of cardiovascular disease during the pandemic, such as unemployment (Dupre et al 2012), inactivity and a possible stressful environment at home.

Although there is evidence on a reduced number of coronary interventions during the pandemic, deaths due to cardiovascular disease increased in the same period in some countries (Del Pinto et al., 2020; Keizman E. et al., 2020), as patients were in a worse condition (Keizman E. et al., 2020). This suggests that patients often preferred to ignore mild symptoms or delay their scheduled examinations to avoid the risk of catching Covid-19 in hospitals. Such delays often lead a deterioration of patients’ clinical condition, leading to severe complications.

To the extent to which such a reduction in operations is a result of delayed treatment, there are serious long-term effects to consider. Delays in treatment for extensive periods, such as being on the surgical waiting list for more than 16 weeks for CABG, contribute to higher mortality and morbidity rates (Da Fonseca et al 2018; Rexius et al 2004; Malaisrie et al 2014). Following a steep decline in cardiac surgery volume in the US, a recent study expects a progressive increase in deferred cases during the pandemic that will require completion within a limited time frame once restrictions ease (Salenger et al 2020). The authors call for early planning by prioritising patients with considerable risks arising from delayed operations (Salenger et al 2020).

This study provides evidence on the short-term impact of the pandemic on cardiac surgery operations. These effects are likely to vary by wave and may extend beyond the end of the pandemic. Future research can study the long-term effects of the pandemic on treatment delays, cardiac surgery operations, cardiovascular-related mortality and cardiovascular health in general.

## Data Availability

Data on each hospital separately cannot be made public without the hospital's consent

## Conflict of interest

None

## Funding

None

## Ethics approval

We used aggregate anonymous data in the study. Nevertheless, ethics approval was given by the Ethics Committee of the Hellenic Society of Thoracic and Cardiovascular Surgeons (26 March 2021, ref 992).

## Checklist

There is no relevant checklist of observational studies.

## Data availability statement

Data on each hospital separately cannot be made public without the hospital’s consent.

## Notes

### Competing Interest Statement

The authors have declared no competing interest.

### Funding Statement

This study did not receive funding

